# Sociodemographic Determinants of Fruit and Vegetable Consumption in Indonesia: Insights from the 2023 Indonesian Health Survey

**DOI:** 10.1101/2025.07.22.25331981

**Authors:** Azimatul Amini, Fitrina Mahardani Kusumaningrum, Digna Niken Purwaningrum, Abdul Wahab, Aditya Lia Ramadona

## Abstract

**Objective:** Inadequate fruit and vegetable consumption is a recognized risk factor for non-communicable diseases. This study aimed to identify the sociodemographic determinants of fruit and vegetable consumption in Indonesia, which would be fundamental to developing targeted public health interventions for non-communicable diseases.

**Methods:** This cross-sectional study utilized secondary data from the 2023 Indonesian Health Survey. We analyzed the data from 595.303 adults aged above 19 years old, comprising variables of fruit and vegetables consumption and sociodemographic determinants. A logistic regression analysis is conducted to examine associations between sociodemographic factors and fruit and vegetable consumption.

**Results:** A significant majority of Indonesians (97.1%) fail to meet the recommended intake of fruits and vegetables. Better fruit and vegetable consumption patterns were observed among females (aOR=1.31; 95% CI=1.24-1.38), individuals with higher education (aOR=1.47; 95% CI=1.23-1.75), those employed (aOR=1.13; 95% CI=1.06-1.20), married (aOR=1.25; 95% CI=1.16-1.34), from lower economic status groups, and residents of Nusa Tenggara (aOR=1.32; 95% CI=1.13-1.53), Maluku (aOR=1.54; 95% CI=1.16-2.06), Papua (aOR=1.95; 95% CI=1.46-2.60), and rural areas (aOR=1.12; 95% CI=1.02-1.23).

**Conclusion:** Indonesians consume very few fruits and vegetables, and this low consumption isn’t uniform. We see significant differences based on factors like sex, education, occupation, marital status, socioeconomic status, and where people live (both urban/rural and specific residential areas). This makes it clear that we urgently need targeted nutrition campaigns and flexible public health policies.

## INTRODUCTION

In 2016, non-communicable diseases (NCDs) accounted for 73% of all deaths in Indonesia. The mortality breakdown consisted of 35% from cardiovascular diseases, 12% from cancer, 6% from diabetes mellitus, and 15% from other NCDs (1). According to the Indonesian Basic Health Research (Riskesdas), the prevalence of NCDs in Indonesia has increased from 2013 to 2018 (2). Prevention of risk factors contributing to NCDs must be prioritized in national health strategies to reduce the future prevalence of these diseases (3).

One of the primary risk factors for NCDs is inadequate consumption of fruits and vegetables (2). NCDs such as obesity are caused by an unhealthy diet (4). Changing unhealthy diets to healthy ones can reduce NCD cases and help people achieve a healthier lifestyle (5). Regular intake of fruits and vegetables can help reduce the risk of cardiovascular diseases, stroke, and hypertension; prevent certain types of cancer such as breast, colon, and lung cancer; and provide protection against neurodegenerative diseases like Alzheimer’s and Parkinson’s (6).

Kucuk (2023), in an analysis of the 2019 Turkish Health Survey, reported that fruit and vegetable consumption was positively associated with multiple sociodemographic and lifestyle variables—such as marital status, educational attainment, active behaviors (cycling and walking), adequate rest, higher income, place of residence, age, and total duration of physical activity (7). While these insights underscore the multifaceted drivers of diet in Turkey, Indonesia’s distinct cultural, economic, and geographic diversity may yield different patterns. Context-specific research is therefore essential to uncover Indonesia’s unique sociodemographic determinants of fruit and vegetable intake.

As non-communicable diseases continue to rise in Indonesia and dietary habits shift alongside rapid urbanization and income growth, the determinants of fruit and vegetable consumption remain underexplored. Addressing this gap is critical for developing finely tuned public health strategies. Accordingly, this study asks: What sociodemographic characteristics are associated with fruit and vegetable consumption in Indonesia, based on the 2023 Indonesian Health Survey (IHS)?

The primary objective of this study is to analyze the association between sociodemographic characteristics and fruit and vegetable consumption in Indonesia using data from the 2023 IHS. In particular, we will (1) characterize national consumption patterns of fruits and vegetables and (2) assess how these patterns vary across key sociodemographic subgroups, thereby informing targeted public health interventions.

## METHODS

### Study Design

This study employed a quantitative approach with a cross-sectional design, utilizing secondary data from the 2023 IHS. The IHS—known in Indonesian as Survei Kesehatan Indonesia (SKI)—is a nationally representative health survey conducted by the Indonesian Ministry of Health (8). The 2023 IHS was selected for this study because it provides the most recent and comprehensive data on individual health status, sociodemographic characteristics, and regional variations. Data collection for the IHS was carried out between August and September 2023.

### Participants and Data Collection

The sample for this study consists of individuals aged 19 years and older, drawn from the 2023 IHS dataset. The independent variables include age, gender, education level, employment status, economic status, marital status, place of residence, and area classification. In the IHS dataset, economic status is assessed based on an index of household ownership of durable goods, which is then categorized into five quintiles. The dependent variable in this study is fruit and vegetable consumption.

Data on fruit and vegetable consumption were collected using the STEPwise instrument developed by the World Health Organization (WHO) (8). This study utilized the individual questionnaire from the 2023 IHS, with instruments sourced from both individual and household questionnaires. These were obtained from the official website of the Ministry of Health of the Republic of Indonesia (www.badankebijakan.kemkes.go.id/hasil-ski-2023/).

The sample included respondents who met the inclusion criteria, which required complete data on all relevant variables: fruit and vegetable consumption patterns, age, sex, education level, employment status, economic status, marital status, place of residence, and area classification. Respondents were categorized into two groups based on their dietary behavior. Those who consumed an adequate amount of fruits and vegetables during the past week were classified as the “healthy group,” while those who did not were classified as the “less healthy group.” Adequate consumption was defined as the intake of at least five servings of fruits and/or vegetables per day, for seven consecutive days (7).

### Statistical Analysis

Respondents under the age of 19 were excluded to align with the study’s inclusion criteria, which focused on individuals aged 19 and above. Initial dataset configurations were performed according to the guidelines outlined in the 2023 IHS Data User Manual. This setup included the application of sampling weights, as the IHS 2023 employed a complex sampling design, ensuring that the analytical results are nationally representative.

Missing data were identified in the economic status variable (n=46). Given the small proportion relative to the total sample size (n=593,303), these cases were excluded from the analysis. Following data cleaning, variables were categorized according to the study’s inclusion criteria.

Descriptive statistical analysis was conducted to summarize the data. Frequencies and percentages were used to describe both independent and dependent variables, with univariate results presented in tabular format. Bivariate analysis was performed using the chi-square test and odds ratios (ORs). The chi-square test assessed the significance of associations between each sociodemographic variable and fruit and vegetable consumption, while ORs measured the strength of these associations. A significance level of p < 0.05 was used for all bivariate tests.

Multivariable analysis was conducted using logistic regression. Only independent variables that demonstrated statistically significant associations in the bivariate analysis were included in the regression model. This approach was intended to minimize the influence of non-significant variables on the model’s outcomes. A Goodness-of-Fit test was performed to evaluate the adequacy of the logistic regression model and ensure its suitability for further inference.

### Ethics Statement

This study received ethical approval from the Medical and Health Research Ethics Committee of the Faculty of Medicine, Public Health, and Nursing, Universitas Gadjah Mada, under permit number KE/FK/0117/EC/2025.

## RESULTS

As presented in Table 1, only 2.9% of respondents met the criteria for a healthy fruit and vegetable consumption pattern. The majority of participants (83.6%) were within the 19–59 age range. The sample was slightly skewed toward female respondents, with most individuals reporting marital status as married and employment status as employed. Educational attainment was predominantly at the primary level, representing 50.2% of the sample. Economic status was distributed relatively evenly across quintiles. Geographically, the largest proportions of respondents resided in Java (28.5%) and Sumatra (31.0%), with smaller distributions across Sulawesi, Kalimantan, Bali, Nusa Tenggara, Maluku, and Papua.

**Table 1.** Description of respondents based on diet and sociodemographic characteristics.

| Variables | N | % |
| --- | --- | --- |
| N | 593.303 | 100,0 |
| **Dietary pattern category** |  |  |
| Unhealthy dietary pattern | 576.119 | 97,1 |
| Healthy dietary pattern | 17.184 | 2,9 |
| **Age** |  |  |
| 19-59 years | 495.964 | 83,6 |
| ≥60 years | 97.339 | 16,4 |
| **Sex** |  |  |
| Male | 268.591 | 45,3 |
| Female | 324.712 | 54,7 |
| **Education level** |  |  |
| No formal education | 30.188 | 5,1 |
| Primary education | 298.020 | 50,2 |
| Secondary education | 192.709 | 32,5 |
| Higher education | 72.386 | 12,2 |
| **Employment status** |  |  |
| Unemployed | 196.292 | 33,1 |
| Employed | 397.011 | 66,9 |
| **Economic status** |  |  |
| Lowest | 112.041 | 20,0 |
| Lower-middle | 113.283 | 20,2 |
| Middle | 110.865 | 19,8 |
| Upper-middle | 111.938 | 20,0 |
| Highest | 112.030 | 20,0 |
| **Marital status** |  |  |
| Unmarried | 124.933 | 21,1 |
| Married | 468.370 | 78,9 |
| **Place of residence** |  |  |
| Jawa | 168.918 | 28,5 |
| Sumatra | 183.951 | 31,0 |
| Kalimantan | 56.103 | 9,5 |
| Sulawesi | 88.832 | 15,0 |
| Bali | 13.730 | 2,3 |
| Nusa Tenggara | 36.100 | 6,1 |
| Maluku | 20.283 | 3,4 |
| Papua | 25.386 | 4,3 |
| **Area classification** |  |  |
| Urban | 317.614 | 53,5 |
| Rural | 275.689 | 46,5 |

Source: IHS 2023

Bivariate analysis (Table 2) indicated statistically significant associations between fruit and vegetable consumption patterns and several sociodemographic variables, including sex, educational attainment, employment status, economic status, marital status, place of residence, and area classification. Age did not demonstrate a significant association and was therefore excluded from subsequent multivariable analysis.

**Table 2.** Results of the Analysis on the Association Between Sociodemographic Factors and Fruit and Vegetable Consumption in Indonesia.

| Variables | Fruit and vegetable diet (%) |  | OR (CI 95%) | aOR (CI 95%) |
| --- | --- | --- | --- | --- |
|  | Less Healthy | Healthy |  |  |
| **Age** |  |  |  |  |
| 19-59 years | 83,6 | 83,1 | *Reference* |  |
| ≥60 years | 16,4 | 16,9 | 1,07 (1,00-1,14) |  |
| **Sex** |  |  |  |  |
| Male | 45,4 | 41,6 | *Reference* | *Reference* |
| Female | 54,6 | 58,4 | 1,23 (1,18-1,30)* | 1,31 (1,24-1,38)* |
| **Education level** |  |  |  |  |
| No formal education | 5,1 | 3,8 | *Reference* | *Reference* |
| Primary education | 50,5 | 41,9 | 1,03 (0,88-1,20) | 0,99 (0,84-1,15) |
| Secondary education | 32,4 | 34,5 | 1,36 (1,16-1,61)* | 1,16 (0,98-1,37) |
| Higher education | 12,0 | 19,8 | 2,29 (1,93-2,72)* | 1,47 (1,23-1,75)* |
| **Employment status** |  |  |  |  |
| Unemployed | 33,1 | 31,9 | *Reference* | *Reference* |
| Employed | 66,9 | 68,1 | 1,06 (1,01-1,11)* | 1,13 (1,06-1,20)* |
| **Economic status** |  |  |  |  |
| Lowest | 19,6 | 32,9 | *Reference* | *Reference* |
| Lower-middle | 20,2 | 20,9 | 0,57 (0,52-0,62)* | 0,62 (0,56-0,68)* |
| Middle | 19,9 | 16,4 | 0,45 (0,41-0,50)* | 0,50 (0,46-0,56)* |
| Upper middle | 20,1 | 15,4 | 0,44 (0,39-0,48)* | 0,49 (0,44-0,54)* |
| Highest | 20,2 | 14,4 | 0,42 (0,37-0,47)* | 0,45 (0,40-0,51)* |
| **Marital status** |  |  |  |  |
| Unmarried | 21,2 | 17,5 | *Reference* | *Reference* |
| Married | 78,8 | 82,5 | 1,29 (1,20-1,39)* | 1,25 (1,16-1,34)* |
| **Place of residence** |  |  |  |  |
| Jawa | 28,3 | 32,7 | *Reference* | *Reference* |
| Sumatra | 31,2 | 24,8 | 0,85 (0,76-0,95)* | 0,85 (0,76-0,95)* |
| Kalimantan | 9,5 | 7,1 | 0,82 (0,71-0,95)* | 0,81 (0,70-0,94)* |
| Sulawesi | 14,9 | 15,8 | 1,05 (0,93-1,18) | 1,07 (0,95-1,22) |
| Bali | 2,3 | 2,1 | 0,93 (0,76-1,14) | 0,81 (0,66-0,99)* |
| Nusa Tenggara | 6,0 | 7,6 | 1,18 (1,02-1,36)* | 1,32 (1,13-1,53)* |
| Maluku | 3,4 | 4,1 | 1,43 (1,10-1,90)* | 1,54 (1,16-2,06)* |
| Papua | 4,2 | 5,8 | 1,95 (1,46-2,60)* | 1,95 (1,46-2,60)* |
| **Area Classification** |  |  |  |  |
| Urban | 53,4 | 59,1 | *Reference* | *Reference* |
| Rural | 46,6 | 40,9 | 0,86 (0,79-0,95)* | 1,12 (1,02-1,23)* |

\*p-value<0,05
Source: IHS 2023

Multivariable logistic regression analysis revealed that sex, higher education, employment status, economic status, marital status, and regional residence remained significantly associated with healthy dietary patterns after adjusting for potential confounders. Female respondents were 1.31 times more likely to exhibit healthy fruit and vegetable consumption compared to male respondents (95% CI: 1.24–1.38). Higher education was positively associated with healthy dietary behavior, with respondents possessing higher education being 1.47 times more likely to meet recommended intake levels compared to those with no formal education (95% CI: 1.23–1.75). Employment status also showed a positive association, with employed individuals being 1.13 times more likely to consume fruits and vegetables adequately than their unemployed counterparts (95% CI: 1.06–1.20).

Contrary to expectations, higher economic status was inversely associated with healthy fruit and vegetable consumption. Married individuals were 1.25 times more likely to adhere to healthy dietary patterns compared to unmarried individuals (95% CI: 1.16–1.34). Regional analysis indicated that respondents residing in Sulawesi, Nusa Tenggara, Maluku, and Papua were more likely to consume fruits and vegetables adequately compared to those in Java. In contrast, respondents from Sumatra, Kalimantan, and Bali exhibited lower odds of healthy consumption. Additionally, rural residence was associated with a higher likelihood of healthy dietary behavior compared to urban residence (aOR = 1.12).

## DISCUSSION

The present analysis revealed that a significant majority of respondents (97.1%) did not meet the recommended daily intake of fruits and vegetables, indicating a pervasive pattern of inadequate dietary behavior within the study population. This finding is critical, as sufficient consumption of fruits and vegetables is a well-established protective factor against various NCDs, including diabetes, chronic respiratory conditions, hearing and visual impairments, asthma, and depression [10]. Globally, low fruit intake has been identified as a major risk factor contributing to both mortality and disability, as evidenced by a systematic analysis of Global Burden of Disease (GBD) data from 195 countries between 1990 and 2017 [11].

Our findings underscore the necessity for health promotion strategies aimed at increasing fruit and vegetable consumption to be specifically tailored to distinct socio demographic groups. Key factors warranting consideration for targeted interventions include sex, education level, employment status, economic status, marital status, place of residence, and area classification. Priority for interventions should be given to individuals who are male, possess lower educational attainment, are unemployed, belong to higher economic status groups, are unmarried, reside in regions such as Sumatra, Kalimantan, or Bali, and live in urban areas.

Gender disparities in dietary behavior are particularly salient, with women consistently demonstrating a higher propensity to consume fruits and vegetables compared to men. This observation aligns with previous studies reporting significantly higher intake levels among women across various contexts [7, 12, 13, 14]. For instance, Li et al. (2022) found that women in China consumed more fruit than men [14], and similar patterns were observed in Turkey, where women exhibited higher fruit and vegetable consumption than their male counterparts [7]. These differences may be partly attributed to traditional gender roles in household food management, wherein women are more frequently involved in purchasing ingredients for daily meals and overseeing household dietary practices, thereby potentially increasing their personal intake of fruits and vegetables.

A consistent positive correlation was observed between higher educational attainment and healthier dietary patterns. This trend is consistent with other research indicating that Source: IHS 2023 education level significantly influences fruit and vegetable consumption [7, 15–17]. Consequently, efforts to enhance fruit and vegetable intake could be prioritized at the individual level, targeting those with no formal education, followed sequentially by individuals with primary, secondary, and higher educational backgrounds. Health professionals are further advised to develop health promotion strategies that are specifically tailored to the educational characteristics of their target groups.

Our study also found that unemployed individuals were less likely to consume fruits and vegetables, a result corroborated by previous research [16, 18]. Employment status can affect fruit and vegetable intake, as individuals in formal employment often have stable incomes and better access to healthy foods, while unemployed individuals may face financial constraints that limit their ability to purchase such foods [7].

Interestingly, the relationship between economic status and fruit and vegetable consumption in this study diverged from previous findings, which typically report higher intake among wealthier individuals [7, 13, 14]. Here, individuals in the highest economic group consumed fewer fruits and vegetables than those in the lowest economic group. This discrepancy may be explained by the measurement of economic status, which in this study was based on ownership of durable goods rather than actual disposable income. Thus, a materially affluent individual may not necessarily translate to a healthier diet, as intake is influenced by factors beyond purchasing power, such as time constraints, personal preferences, social environment, and lifestyle choices. This inverse association between higher economic status (as measured by durable goods) and lower fruit/vegetable intake highlights a modern lifestyle trend that warrants further investigation to better understand consumer behavior.

Married individuals exhibited a greater tendency to consume fruits and vegetables compared to their unmarried counterparts, a finding consistent with previous research showing higher intake among married populations [7, 16, 19]. Marital status therefore warrants consideration as a determinant in the design of community nutrition interventions, as the presence of a spouse can serve as a supportive factor for fruit and vegetable consumption. Health promotion interventions can guide individuals to strengthen household support mechanisms to establish sustainable fruit and vegetable consumption behaviors.

Geographical disparities in fruit and vegetable consumption were also evident, with regions such as Sulawesi, Nusa Tenggara, Maluku, and Papua demonstrating higher proportions of healthy diets compared to Java. This suggests that less developed areas like Nusa Tenggara and Papua consume more fruits and vegetables than more developed regions such as Sumatra, Kalimantan, and Java [15]. Place of residence influences eating patterns through factors such as access to healthy food, prevailing community habits, and local food availability. These findings emphasize the critical need for context-specific interventions that are tailored to regional and population characteristics. Policymakers should leverage evidence-based data to formulate targeted strategies aimed at reducing dietary disparities across regions and social strata.

Within the framework of the Social Determinants of Health (SDH), sociodemographic factors—including gender, education, occupation, economic status, and geographic location—act as social determinants that can profoundly influence individual health behaviors, such as fruit and vegetable consumption. Discrepancies in sociodemographic characteristics, access to resources, health knowledge, and social environments collectively illustrate how these social determinants contribute to disparities in public health outcomes. Consequently, the findings of this study provide a foundational basis for designing multi-level, equity-oriented intervention strategies.

Given that inadequate fruit and vegetable consumption is a well-established risk factor for NCDs, dedicated attention is required to promote healthier diets across all segments of society. This aligns with recommendations from other studies, which identify the promotion of fruit and vegetable intake as a pivotal strategy in the prevention and control of NCDs. Furthermore, these promotional efforts should extend to individuals already living with NCDs, ensuring they achieve the recommended fruit and vegetable intake as advised by government guidelines [10].

To foster improved dietary patterns, particularly in terms of fruit and vegetable consumption, multi-level interventions are indispensable. These interventions should encompass educational initiatives, environmental modifications, and supportive public policies. Such efforts should be implemented gradually, coherently, and sustainably, as recommended by Woodside et al. (2023). By integrating individual, community, and structural approaches, it is anticipated that the prevalence of low fruit and vegetable intake in Indonesia can be minimized, thereby contributing to a healthier and more equitable society.

The study identified a particularly vulnerable group with the lowest fruit and vegetable intake, comprising males, individuals without formal education, the unemployed, those in higher economic strata (as measured by durable goods), unmarried individuals, and residents of Sumatra, Kalimantan, Bali, and urban areas. Their heightened vulnerability stems from limited access, lower motivation, reduced awareness, specific social roles, and insufficient environmental support. Addressing these disparities necessitates robust multi-sectoral collaboration. For instance, the Ministry of Agriculture could facilitate the development of community gardens or affordable fruit stalls. The National Food Agency could provide fresh vegetable packages to low-income families. The Ministry of Manpower and private sectors could integrate nutrition education into labor programs. The Ministry of Higher Education, Science, and Technology should engage local leaders as change agents in nutrition education. Health promotion campaigns can be bolstered through peer support and the involvement of male role models who actively practice healthy eating, perhaps through culturally sensitive campaigns that resonate with the target demographic.

The government plays a crucial role in supporting fruit and vegetable consumption through price regulations and market governance. To incentivize healthy eating, policies could include subsidizing local horticultural products, taxing highly processed foods, and regulating healthy food distribution in public institutions. These policy interventions are essential to counteract prevailing market trends that often favor inexpensive, calorie-dense, and nutrient-poor foods [21,22]. Therefore, beyond educational initiatives, policy-driven structural strategies are paramount to ensuring equitable and sustainable health promotion outcomes.

The measurement of economic status using household durable goods ownership in the IHS dataset revealed a counterintuitive association: higher economic status correlated with lower fruit and vegetable consumption. This type of measurement, based on static assets, may not accurately reflect an individual’s current, fluid purchasing ability or disposable income for daily dietary choices. This suggests that increased wealth, when assessed by durable goods, does not automatically translate to healthier diets, as intake is influenced by a complex interplay of factors beyond purchasing power, including time constraints, personal preferences, social environment, and modern lifestyle choices. This specific modern lifestyle trend merits further dedicated research to gain a deeper understanding of consumer behavior.

When promoting fruit and vegetable consumption, it is crucial to transition from generalized, equality-based methods to targeted, equity-focused approaches. These approaches must address the unique needs of vulnerable groups. For example, unemployed or unmarried men might benefit significantly from visual education, peer support networks, and readily available, affordable fruits and vegetables. Individuals with lower economic status could be supported through direct subsidies or the establishment of community fruit stalls. An equity-based approach ensures that interventions are more suitable, precisely targeted, and ultimately more effective in reducing consumption disparities [23].

Finally, observations from Maluku underscore that fruit and vegetable consumption is shaped by a confluence of individual, social, and spatial factors. In rural Maluku, many households possess fruit trees (e.g., mangoes), providing direct and immediate access to fruits [24]. This contrasts with urban areas, where limited land availability reduces local food sources and increases reliance on market purchases. Our findings indicate higher consumption in rural areas, while urban areas showed no significant association. This highlights the imperative to consider spatial and ecological factors in nutrition policies, advocating for initiatives such as supporting urban farming, promoting home gardens, and revitalizing traditional markets to boost fruit and vegetable intake in urban settings.

## Data Availability

All data generated in this study are accessible upon request via the following link: badankebijakan.kemkes.go.id/hasil-ski-2023/

## NOTES

## Acknowledgements

None.

